# Graphical Learning and Causal Inference for Drug Repurposing

**DOI:** 10.1101/2023.07.29.23293346

**Authors:** Tao Xu, Jinying Zhao, Momiao Xiong

## Abstract

Gene expression profiles that connect drug perturbations, disease gene expression signatures, and clinical data are important for discovering potential drug repurposing indications. However, the current approach to gene expression reversal has several limitations. First, most methods focus on validating the reversal expression of individual genes. Second, there is a lack of causal approaches for identifying drug repurposing candidates. Third, few methods for passing and summarizing information on a graph have been used for drug repurposing analysis, with classical network propagation and gene set enrichment analysis being the most common. Fourth, there is a lack of graph-valued association analysis, with current approaches using real-valued association analysis one gene at a time to reverse abnormal gene expressions to normal gene expressions.

To overcome these limitations, we propose a novel causal inference and graph neural network (GNN)-based framework for identifying drug repurposing candidates. We formulated a causal network as a continuous constrained optimization problem and developed a new algorithm for reconstructing large-scale causal networks of up to 1,000 nodes. We conducted large-scale simulations that demonstrated good false positive and false negative rates.

To aggregate and summarize information on both nodes and structure from the spatial domain of the causal network, we used directed acyclic graph neural networks (DAGNN). We also developed a new method for graph regression in which both dependent and independent variables are graphs. We used graph regression to measure the degree to which drugs reverse altered gene expressions of disease to normal levels and to select potential drug repurposing candidates.

To illustrate the application of our proposed methods for drug repurposing, we applied them to phase I and II L1000 connectivity map perturbational profiles from the Broad Institute LINCS, which consist of gene-expression profiles for thousands of perturbagens at a variety of time points, doses, and cell lines, as well as disease gene expression data under-expressed and over-expressed in response to SARS-CoV-2.

## Introduction

The path to drug development involves long processing: basic research, identification of the target compound, hit identification and validation, hit-to-lead optimization, drug candidate selection, preclinical development, three elaborate and lengthy phases of human clinical trials, regulatory review and approval, and post-market monitoring^1^. New drugs often take 9.5-15 years to get into the market at a cost as high as $2.6 billion with only a fraction of the drugs getting to market^2^. To address these issues, drug repurposing strategies which can take existing drugs approved by FDR to safely treat disease, other compounds that have been studied as potential drugs, or advancing previously studied but unapproved drugs, are used to find new medical indications for these old compounds^3,4^. The advantages of re-purposing drugs are that they can be faster, cheaper, less risky and have higher success rates getting to market than general drug discovery approaches^3^.

The major task of drug repurposing is to discover new associations of drugs with diseases^5^. The methods for drug repurposing include (1) genetic association analysis for phenotype screening^6,7^, (2) target discovery using bioinformatics and artificial intelligence^9^, (3) systems biology approaches for identifying new drug targets or repurposing opportunities^10^, (4) use drug-repurposing databases such as “Connectivity Map”^11^ and “LINCS”^12^ which provide a resource for researchers to identify new uses for existing drugs by integrated analysis of the gene expression profiles of drug-treated cells and disease-affected cells and (5) using clinical data from electronic health records (EHRs) to predict drug response in humans for many repurposing^4,13^.

Gene expression profiles which can connect drug perturbation, disease gene expression signatures and clinical data are valuable resources for discovering potential drug repurposing indications^4,14^. A popular approach to using drug-induced and disease-specific gene expression data for identification of potential repurposed drugs is investigation of the ability of the drug to reverse over-gene expressions and under-gene expressions in the disease samples to normal gene expressions in the control samples^14,15^.

However, the current gene expression reversal approach has several limitations. First limitation is that most gene expression reversal methods focus on validation of reversal expression of individual genes. Drug efficacy is determined based on the ability to reverse altered expression of individual genes^4,14,15^. Drug often induce many gene expressions and disease is linked to abnormal expression of a quite large number of genes which are not regulated properly in the disease. A large number of drug induced gene expressions and disease specific abnormal gene expressions are interconnected and form complex networks. Therefore, individual gene-based approaches for identifying repurposable drugs are not efficient.

Second limitation is a lack of causal approaches to identifying drug-repurposing candidates. There are some works on reconstruction of drug-induced gene expression networks and disease specific gene expression networks and their integrations for drug repurposing analysis. However, these reconstructed networks are undirected networks. Very few methods have been developed for reconstruction of large causal gene expression networks. There has also been a lack of causal network approaches to search for drug-repurposing candidates.

Third limitation is that very few methods for information passing and summarizing on a graph have been used for drug repurposing analysis. Most approaches for massage passing and summarizing are classical network propagation and gene set enrichment analysis^16^.

Fourth limitation is a lack of graph-valued association analysis. The current approaches to reversing abnormal gene expressions to normal gene expressions use real valued association analysis, one gene association analysis at a time^17^.

To overcome these limitations, we propose a causal inference and graph neural network – based framework for identification of drug-repurposing candidates. The classical methods for learning causal networks are often formulated as a discrete optimization problem where the search space of causal networks is combinatorial, which seriously limits the size of reconstructed causal networks^18^. Zheng et al^19^. formulated a causal network as a structural equation model (SEM), acyclic constraint in term of smooth continuous function and hence reconstruction of the causal network as a continuous optimization problem, avoiding combinatorial formulization. Unfortunately, their formulation considered only endogenous variables. In drug induced gene expression causal networks, gene expressions are taken as endogenous variables and dosages of drugs are taken as exogenous variables. We extend the SEM with only endogenous variables to the SEM with both endogenous and exogenous variables^20,21^.

We use directed acyclic graph neural networks (DAGNN) to aggregates and summarize information from the spatial domain of causal network with no features for nodes and edges which are called non-attributed^22-24^. Thus, DAGNNs that use both the graph structure and node features produce a real number representation which is called summary statistic for DAGNN. We calculate representations of DAGNNs for drug’s perturbated gene expression causal network and disease’s perturbated gene expression causal networks. Finally, to measure degree of reversing altered gene expressions of disease to normal gene expressions by drugs, we develop graph regression. Specifically, the representation of DAGNN for causal gene expression network of disease is regressed on the representation of DAGNN for causal gene expression networks perturbated by drugs to select the candidate drugs^25,26^.

The proposed methods for drug repurposing are applied to phase I and II L1000 connectivity map perturbational profiles from Broad Institute LINCS center under access code GSE70138 and GSE92742 which consist of gene-expression profiles for thousands of perturbagens at a variety of time points, doses, and cell lines^27, 28^, and under-expressed and over-expressed responses to SARS-CoV-2 with access code GSE147507^29,30^.

## Results

### Outline of the computational pipeline for identifying drug repurposing candidates

We are developing a novel approach for identifying drug repurposing candidates that combines graph learning and causation analysis. A schematic of the computational pipeline is depicted in Figure 1. Our approach begins by collecting drug perturbated gene expression profiles from publicly available resources such as Connectivity Map (CMap) and LINCS. We then reconstruct large-scale drug-induced causal networks from these gene expression profiles by formulating graph-based causal inference as a continuous optimization problem to learn directed acyclic graphs (DAGs) (Methods). To assess the potential of a drug to reverse disease gene expressions, we utilize graph neural networks (GNNs) to summarize information in the drug-induced causal networks.

**Figure 1.**
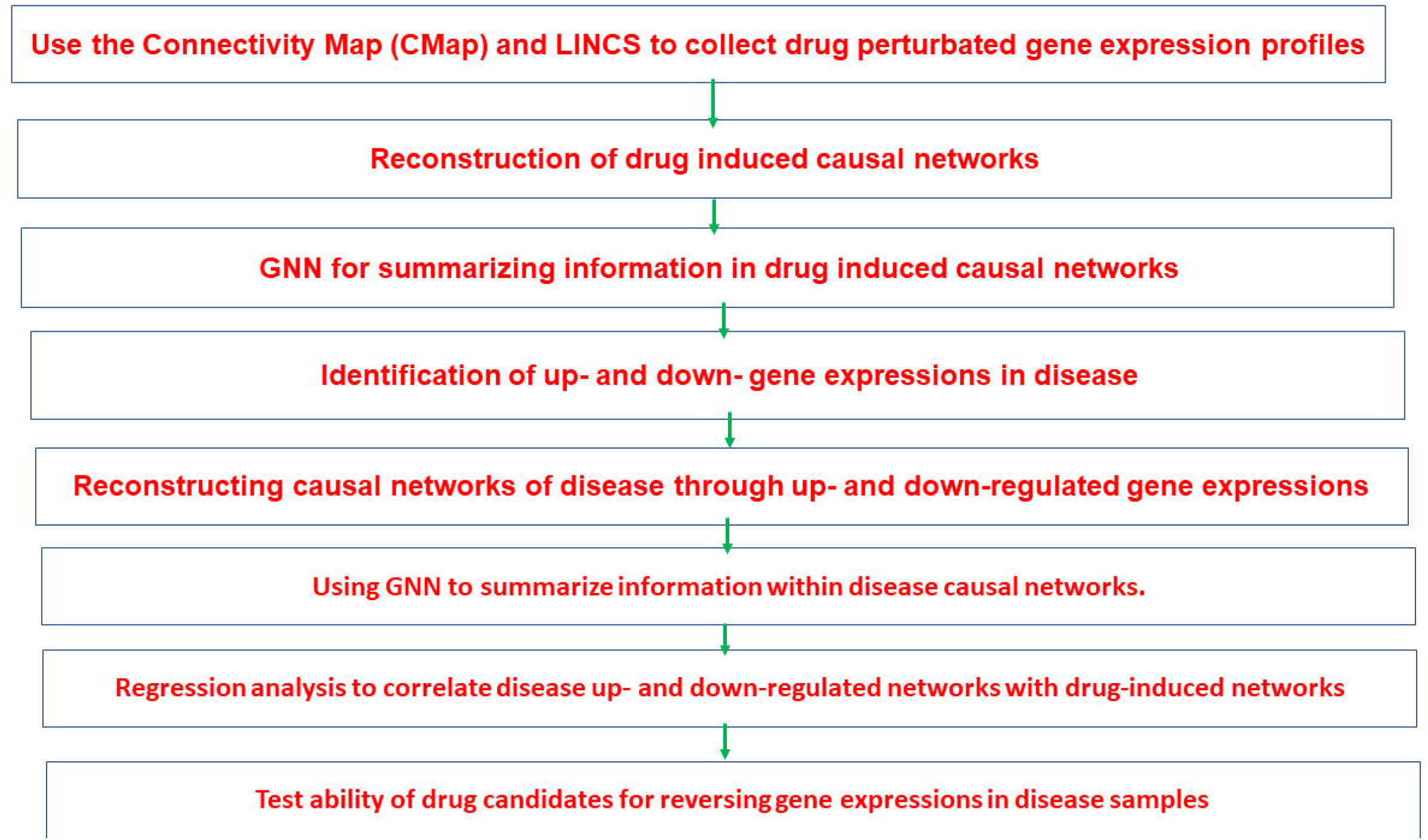
Outline of the computational pipeline for identifying drug repurposing candidates.

The next step is to identify the up- and down-regulated gene expressions in the disease and construct causal networks using the same methods employed for reconstructing drug-induced causal networks. Once we have reconstructed the causal up- and down-expression networks, we will summarize their information using GNNs.

To assess the ability of drug candidates to reverse gene expressions in disease samples, we will perform a regression analysis of disease up- and down-regulated networks on drug-induced networks. A negative coefficient obtained from regressing the disease up-regulated network on the drug-induced network will indicate that the drug can reverse up-regulated gene expression to normal levels. Similarly, a positive coefficient obtained from regressing the disease down-regulated network on the drug-induced network will indicate that the drug can reverse disease down-regulated gene expressions to normal level.

Finally, we will evaluate the ability of drug candidates to reverse gene expressions in disease samples through the regression analysis described above. This approach has the potential to significantly accelerate drug discovery and repurposing efforts.

### Reconstruction of causal networks and simulations

Gene expression causal networks are modeled as DAGs where each node denotes either endogenous gene expression variable or exogenous drug variable and each edge denotes either regulatory relationship between endogenous variables or perturbation of exogenous drug variable to the endogenous gene expression variables (Methods). The classical methods for inferring DAGs are often formulated as a discrete optimization problem where we require a large number of search^19^. The intractable search space seriously limit the size of reconstructed DAG. To overcome combinatorial optimization limitation and to make the DAG search tractable, Zheng et al^19^ formulate DAG learning as a continuous optimization problem with acyclic constraints and least square loss function. The widely used gradient and non-smooth optimization methods can be efficiently used to solve continuous optimization problem^31,32^. Zheng et al.’s approach to learning DAGs considers only the endogenous variables. However, causal gene expression networks for drug repurposing need to consider both endogenous and exogenous variables.

Therefore, we extend Zheng et al.’s approach to including both endogenous and exogenous variables in the DAGs. Specifically, we consider the structural equation model (SEM) (Methods):

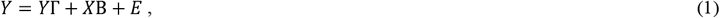

where *Y* is a matrix of observed endogenous variable value (gene expression levels), *X* is a matrix of observed exogenous variable values (drug dosages), Γ is a weight matrix describing connections between endogenous variables (gene regulation matrix), *B* is a weight matrix describing the contributions of the exogenous variables to the endogenous variables (drugs to gene expressions, *E* is a matrix of errors.

To evaluate the performance of the proposed causal network analysis, we conducted a series of simulation studies to compare the detection power and false discovery rate (FDR). Simulations consisted of two parts. The first part was to compare with three classical methods: (1) the graphical lasso (GLAAS) method, (2) structural equation model (SEM) and structural equation model coupled with integer programming (SEMIP). The second part was to compare with GLASS for undirected network. The proposed method was referred to continuous optimization method (COM). In all simulations, we assumed the density of edges of 0.05 and density of exogeneous variable of 0.15.

In the first part, we randomly generated 1,000 directed acyclic graphs (networks) with 40 nodes. The simulation results were summarized in Table 1 where directed graphs were converted to undirected graphs when the GLASS was used for simulations. Simulations were carried out for 500 and 1,000 samples. We observed that in all cases the COM had the smallest FDR. Although the COM had the second largest power for 1,000 samples, the SEM that had the largest power for 1,000 samples also had 46% intolerable FDR.

**Table 1.** Power and false discovery rate (FDR) for construction of network with 40 nodes.

| Method | Power | FDR | Number of variates | Number of samples |
| --- | --- | --- | --- | --- |
| SEM | 80 | 35 | 40 | 500 |
| SEMIP | 76 | 32 | 40 | 500 |
| COM | 82 | 32 | 40 | 500 |
| GLASS | 75 | 39 | 40 | 500 |
| SEM | 95 | 46 | 40 | 1000 |
| SEMIP | 82 | 34 | 40 | 1000 |
| COM | 88 | 23 | 40 | 1000 |
| GLASS | 84 | 44 | 40 | 1000 |

In the second part, we increased the number of nodes in the directed acyclic graphs to 200. We used 1,000, 2,000 and 5,000 samples in simulations. All directed graphs were converted to undirected graphs when the GLASS was used for simulations. The simulation results were summarized in Table 2 where for the convenience of comparisons, the FDR for both COM and GLASS were enforced to have the same rates. We surprisingly observed that the power of detecting the true directed acyclic graphs using the COM was much higher than the power of detecting undirected graphs using the GLASS.

**Table 2.** Power and false discovery rate (FDR) for construction of causal network with 200 nodes.

| Performance | N=1000 | N=2000 | N=5000 |
| --- | --- | --- | --- |
| FDR | 0.19 | 0.17 | 0.11 |
| Power | 0.59 | 0.71 | 0.74 |
|  | N=1000 | N=2000 | N=5000 |
| FDR | 0.19 | 0.17 | 0.11 |
| Power | 0.35 | 0.4 | 0.39 |

### Up-expressed and down-expressed regulatory networks in COVID-19 samples

A classical approach to drug repurposing is to evaluate the ability of the candidate drug to reverse alternative under gene expressions or over gene expressions to normal gene expressions. It starts with identifying a set of under-expressed and over-expressed genes in COVID-19 samples. We selected 86 down-expressed genes and 93 up-expressed genes in response to SARS-CoV-2 with access code GSE147507^29,30^, where gene expression profiling and differential expression analysis were performed for post-mortem lung samples from COVID-19-positive patients with biopsied healthy lung tissue from uninfected individuals. The classical methods for assessing the ability of the drug to reverse the alternative gene expression are using either one by one comparisons or cluster by cluster comparison, which ignores the regulatory relations between genes. To evaluate the ability of the drug more accurately for reversing the alternative gene expressions, we reconstructed down-expressed (Figure 2) and up-expressed causal networks (Figure 3). The under-expressed causal network had four hub genes: Eukaryotic Translation Initiation Factor 3 Subunit A (EIF3A), Regulatory Factor X7 (RFX7), Impact RWD Domain Protein (IMPACT) and Polypeptide N-Acetylgalactosaminyltransferase 7 (GALNT7). The over-expressed causal networks had 6 hub genes: dual specificity phosphatase 1(DUSP1), TIR domain containing adaptor molecule 1(TICAM1), Zinc Finger Protein 613 (ZNF613), TSC22 domain family member 2 (TSC22D2), TNF alpha induced protein 3 (TNFAIP3), and retinoic acid receptor responder 2 (RARRES2).

**Figure 2.**
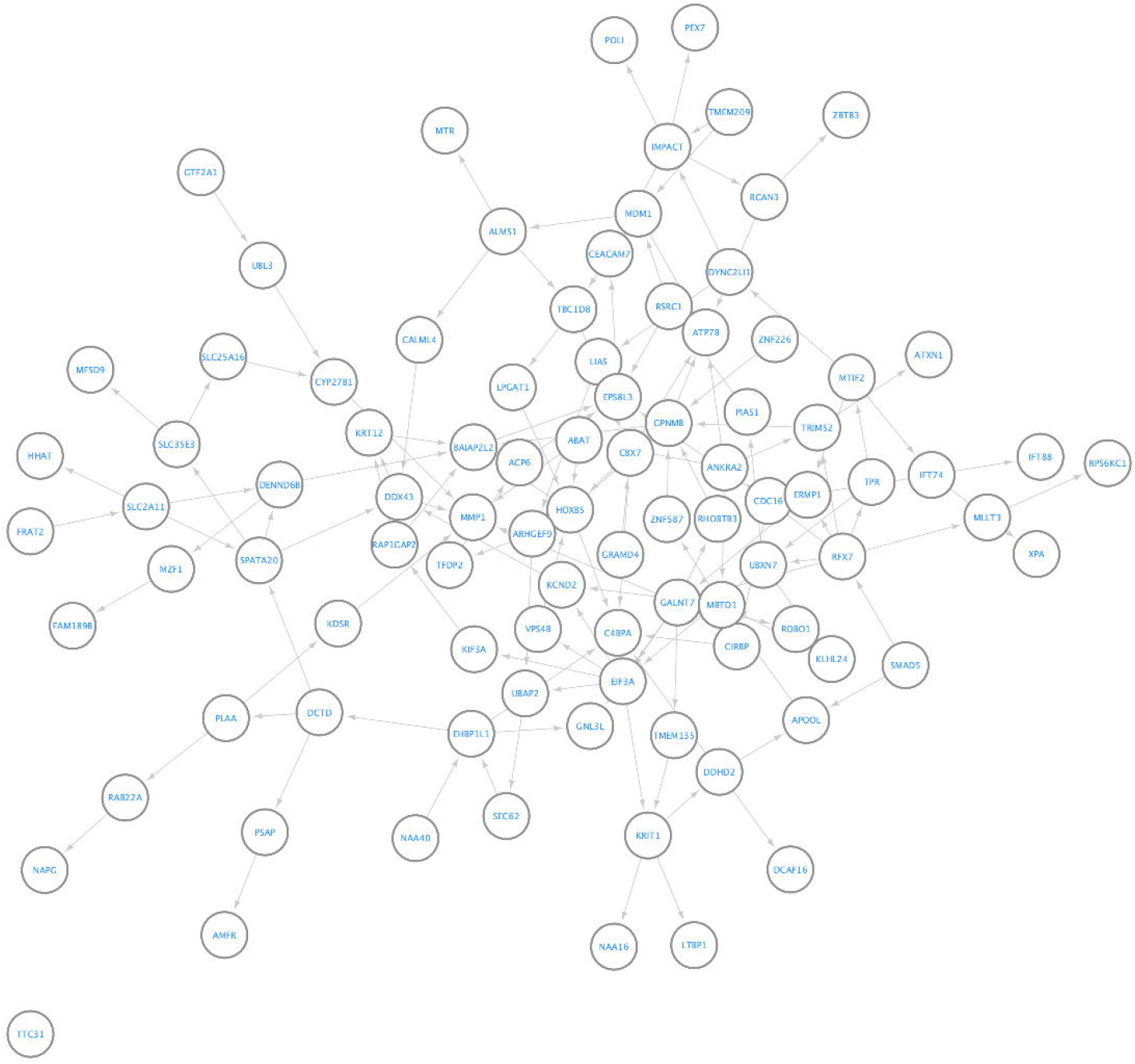
Network of down-regulated genes associated with diseases.

**Figure 3.**
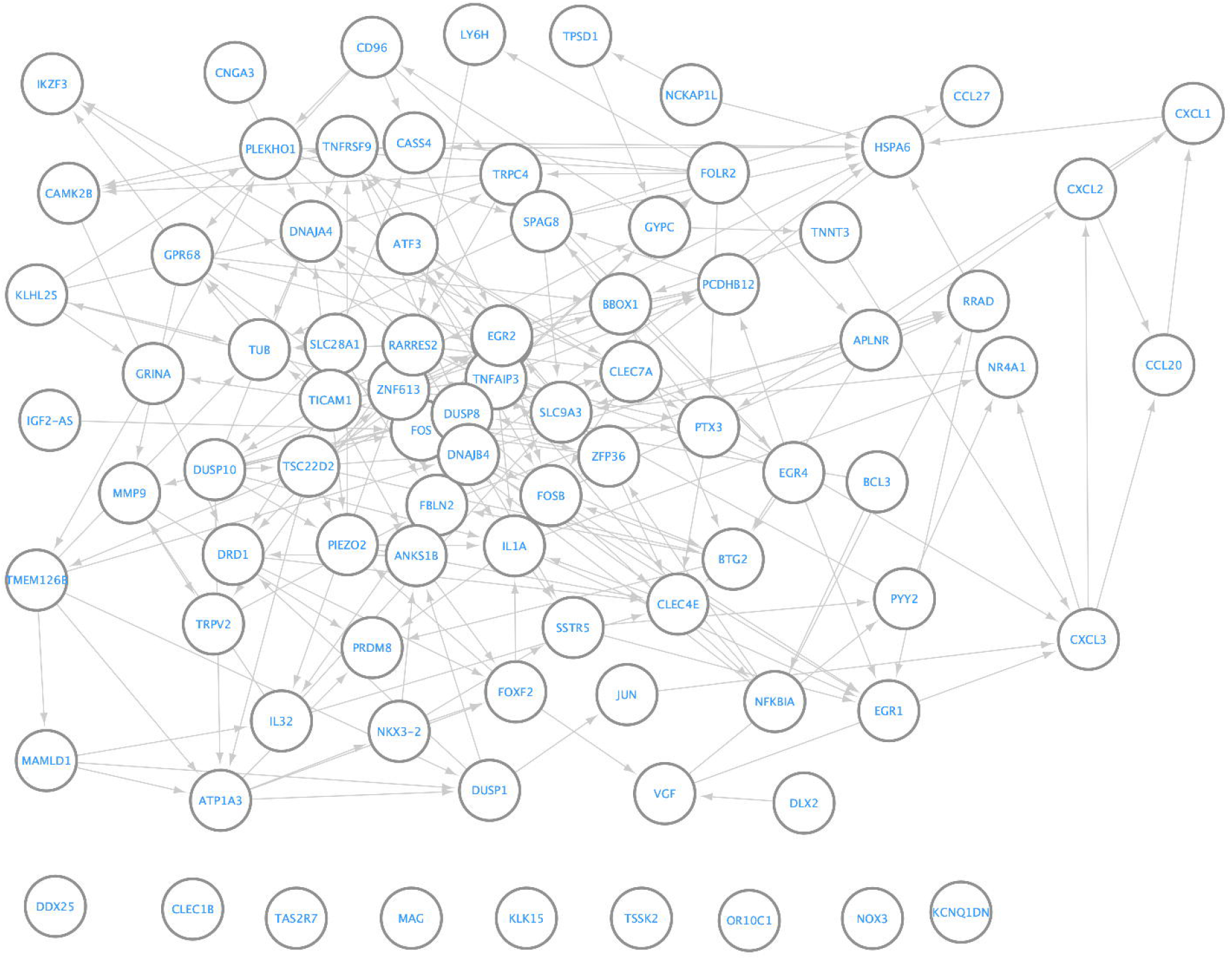
Network of up-regulated genes associated with diseases.

The virus SARS-CoV-2, uses its nonstructural protein 1 (Nsp1) to inhibit translation in cellular, but not viral, through competing with the EIF3A complex for binding to the 40S ribosomal subunit, which prevent loading the mRNA from the host cell for starting translation^33^.

One of the major pathways that RFX7 is involved in is the ciliary signaling pathway^34^. There have been studies that suggest that ciliary dysfunction may contribute to the pathogenesis of COVID-19^35^. Impact plays the role in dysregulation of mitochondrial function which may cause COVID-19^36^. Gene GALNT7 is involved in the O-glycosylation pathway. The surface glycan is involved in viral entry, infection, transmission, antigen, antibody responses, and disease progression which implies that Gene GALNT7 plays an important role in pathogenesis and therapy of COVID-19^37^.

In the over-expressed causal networks, we also observed hub-genes that play an important role in pathogenesis of COVID-19. The DUSP10/MKP5 protein was upregulated in lung tissue samples from patients with severe COVID-19 compared to non-COVID-19 controls^38^, and has been implicated in the MAPK/ERK, JNK and p38 pathways^39^, which are associated with virus infection^40^. TICAM1 plays a critical role in the innate immune response by activating the Toll-like receptor (TLR) signaling pathway, NF-kB pathway and Apoptosis pathways^41^ which lead to the production of pro-inflammatory cytokines and interferons in response to viral infections and plays the role of Immunogenetics in COVID-19^42^. Gene ZNF613 is involved in multiple signaling pathways and cellular processes, including Wnt signaling, cell cycle regulation, TGF-β signaling, and epigenetic regulation. TSC22D2 is involved in the TGF-β, MAPK, P53, Glucocorticoid signaling pathways which regulate cell growth, differentiation, and survival. It has been shown to be associated with COVID19^43^. TNFAIP3 is a negative regulator of the NF-κB pathway. It functions to inhibit the activation of NF-κB^44^. TNFAIP3 also inhibits the activity of other pro-inflammatory signaling pathways such as MAPK and IRF3^45^. SARS-CoV-2 early infection signature identifies TNFAIP3 as one of potential key infection mechanisms and drug targets^46^. RARRES2 is involved in the chemokine, adipokine, MAPK and PI3K/Akt signaling pathways, resulting in the activation and migration of immune cells. and regulates the differentiation and activation of immune cells^47^. It was reported that RARRES2 levels were higher in the serum of COVID-19 patients compared to healthy controls, and that RARRES2 levels were an independent risk factor of mortality^48^. We observed that TICAM1 and TNFAIP3 were involved in NF-kB pathway, TNFAIP3, DUSP10, RARRES2, and TSC22D2 were involved in MAPK pathway, and ZNF613 and TSC22D2 were involved in TGF-β pathway.

### Drug induced causal networks and targets

To identify potential drug targets, we reconstructed drug induced causal networks using phase I and II L1000 perturbational profiles of 978 genes from Broad Institute LINCS center under access code GSE70138 and GSE92742^28^ and 4 drugs: ritonavir, chloroquine, ruxolinib and ribavirin. The number of samples used for drugs ritonavir, chloroquine, ruxolinib and ribavirin, was 149, 232, 204 and 187, respectively. Perturbated gene expressions were taken as endogenous variables and dosages of drug were taken as exogenous variables. The reconstructed causal networks for drugs chloroquine, ritonavir, ruxolinib and ribavirin were presented in Figures 4, 5, Figures S1 and S2, respectively.

**Figure 4.**
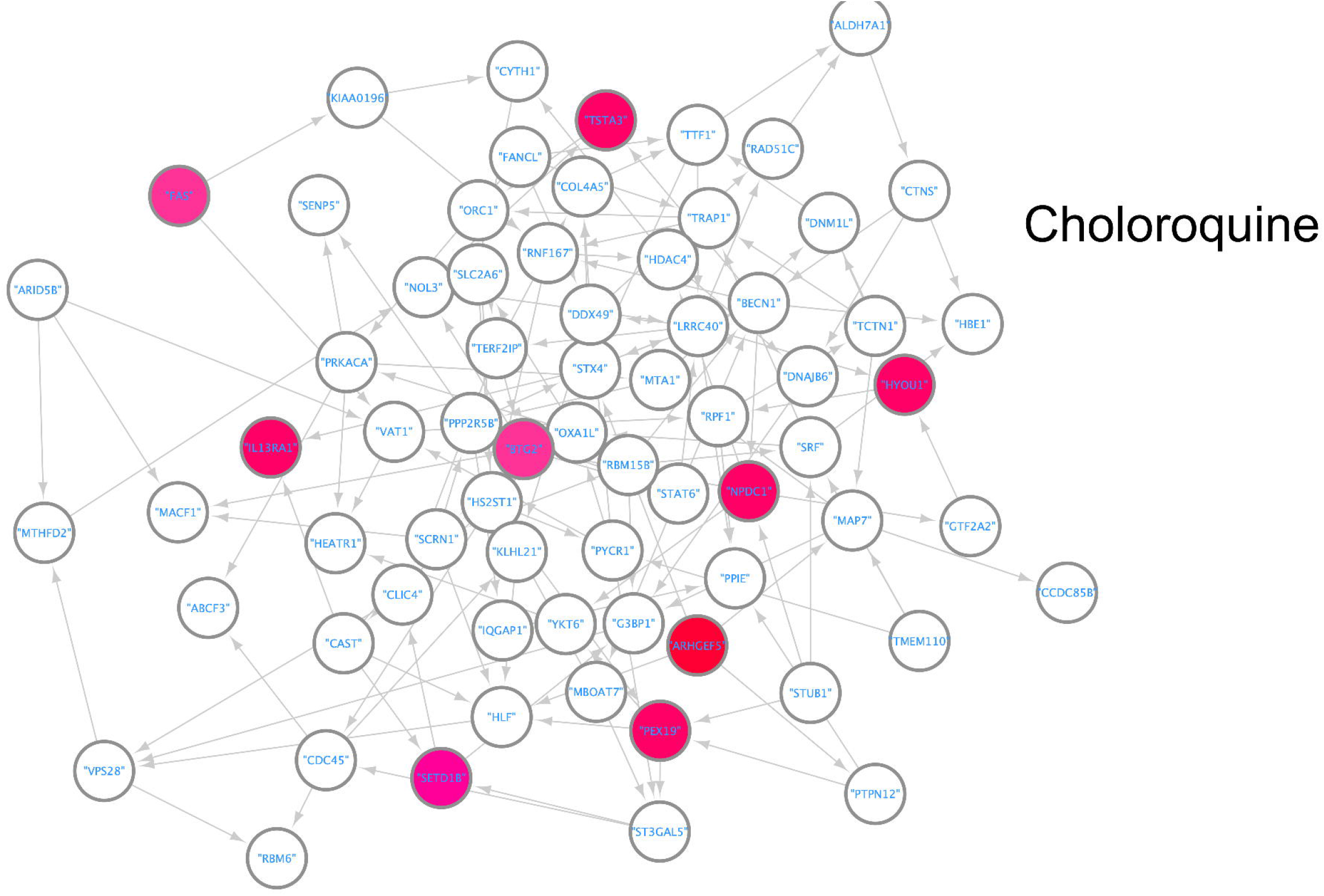
Network of gene expressions induced by Chloroquine with causal implications.

**Figure 5.**
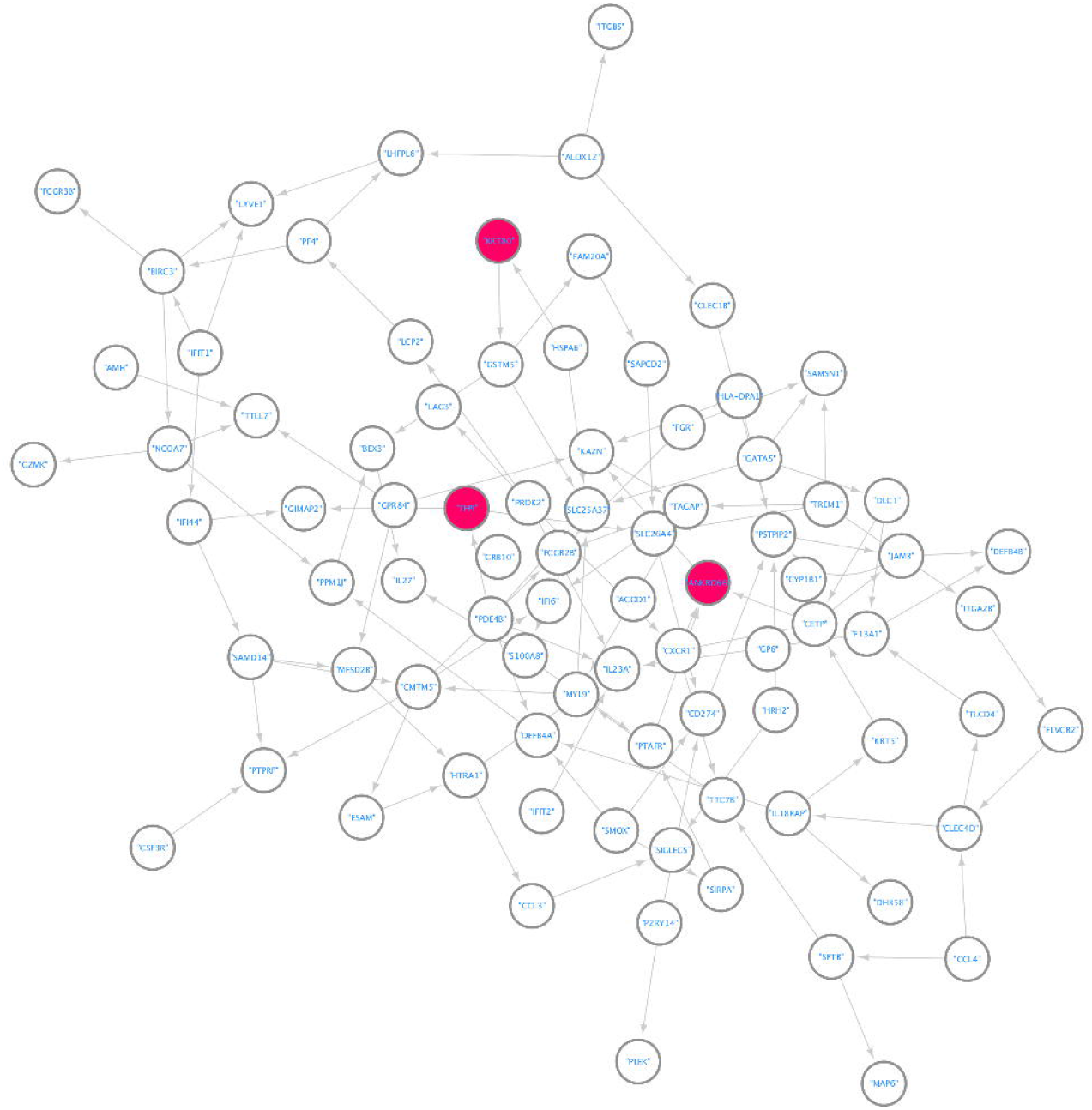
Network of gene expressions induced by Ritonavir with causal implications

Chloroquine is a drug that has been used for the prevention and treatment of malaria, as well as for the treatment of certain autoimmune diseases such as rheumatoid arthritis and lupus. Chloroquine has been shown to have immunomodulatory effects, including the inhibition of cytokine production and the suppression of T-cell activation. Figure 4 showed that the drug chloroquine potentially targeted 9 genes: Fas Cell Surface Death Receptor (FAS), interleukin 13 receptor subunit alpha 1(IL13RA1), SET domain containing 1B, histone lysine methyltransferase (SETD1B), GDP-L-fucose synthase (TSTA3), BTG Anti-Proliferation Factor 2 (BTG2), hypoxia up-regulated 1 (HYOU1), NPC intracellular cholesterol transporter 1 (NPOC1), Rho guanine nucleotide exchange factor 5 (ARHGEF5) and peroxisomal biogenesis factor 19 (PEX19). FAS is involved in the extrinsic apoptotic pathway^49^. Dysregulation of this pathway has been implicated in various diseases, including cancer and autoimmune disorders. Drugs that target this pathway are being developed as potential cancer therapeutics^50^. IL13RA1 is a key player in the immune system and has been identified as a potential drug target for various diseases, including asthma, chronic obstructive pulmonary disease (COPD), and cancer^51^. BTG2 is a protein that has been implicated in various cellular processes. Small molecules that can modulate the activity of BTG2 could potentially be developed into drugs that could be used to treat cancer or other diseases^52^. HYOU1 has been implicated in various diseases, including cancer, neurodegenerative diseases, and cardiovascular diseases and is a potential therapeutic target for cancer^53^. ARHGEF5 plays a role in the regulation of cytoskeletal dynamics and is involved in various cellular processes such as cell migration, adhesion, and proliferation^54^. Studies have shown that targeting ARHGEF5 may inhibit tumor growth and invasion^55^. In addition, ARHGEF5 has been implicated in the development of inflammatory diseases such as asthma and rheumatoid arthritis. Mutations in PEX19 are associated with a range of human diseases, including Zellweger syndrome and other peroxisome biogenesis disorders^56^. Studies have shown that inhibiting PEX19 can inhibit tumor growth and increase sensitivity to chemotherapy. This suggests that targeting PEX19 could also be a potential therapeutic approach for these diseases^57^.

Ritonavir is an antiretroviral medication that is primarily used to treat HIV (human immunodeficiency virus) infection^58^. Recently, ritonavir has also been used to treat other viral infections, such as hepatitis C and coronavirus including SARS-CoV-2^59^. Figure 5 showed that the drug ritonavir had three target genes: tissue factor pathway inhibitor (TFP1/TRAP), keratin 80 (KRT80) and ankyrin repeat domain 66 (ANKRD66). TFP1/TRAP1 is a member of the heat shock protein 90 (HSP90) family, which is involved in regulating the folding and stability of various proteins. In cancer cells, TFP1/TRAP1 has been found to be upregulated and associated with tumor progression and drug resistance^60^. Several small molecule inhibitors of TFP1/TRAP1 have been developed and tested in preclinical studies, obtaining positive results and suggesting that TFP1/TRAP1 could be a potential therapeutic target for cancer, including breast, lung, and pancreatic cancer^61^. ANKRD66 has a high likelihood to interact with GABRP that is identified as a promising drug target for colorectal cancer^62^. Some studies show that miR-155 represents as a potential target for treating psoriatic skin lesions while KRT80 is a target gene of miR-155^63^.

Ruxolitinib is a medication that is primarily used to treat certain types of blood cancers and bone marrow disorders^64^. In addition, some studies have also shown that ruxolitinib has potential in treating other diseases such as graft-versus-host disease (GVHD) and certain types of lymphomas^65^. Figure S1 showed that the drug ruxolitinib had seven target genes: TERF2 interacting protein (TERF21P), Clathrin Light Chain B (CLTB), ribonuclease/angiogenin inhibitor 1 (RNH1), testis expressed 10 (TEX10), ubiquitin conjugating enzyme E2 A (UBE2A), RAB11, family interacting protein 2 (RAB11FIP2), Eukaryotic Translation Initiation Factor 1B (EIF1B). Among them, TERF2IP is a potential target gene for cancer, cardiovascular diseases, and neurological disorders^66^, TEX10 has been shown to play a role in cell growth and proliferation, making it a potential target for cancer therapies^67^, UBE2A-related XLID is linked to intellectual disability^68^, RAB11FIP2 has played a crucial role in maintaining the structure and function of neuronal synapses^69^ and EIF1B is a protein that plays a role in the initiation of protein synthesis in eukaryotic cells. It has been implicated in various cellular processes, including viral infection, cell growth, and cancer^70^.

Ribavirin is an antiviral drug that targets viral replication by inhibiting viral RNA synthesis and leads to the eventual clearance of the infection. Therefore, ribavirin has been used to treat a variety of viral infections, including hepatitis C virus (HCV) and respiratory syncytial virus (RSV)^71-72^.

We found that a total of ten genes: protein kinase cAMP-activated catalytic subunit alpha (PRKACA), hypoxia up-regulated 1(HYOU1), DnaJ heat shock protein family (Hsp40) member C15 (DNAJC15), integrin linked kinase (ILK), TNFAIP3 interacting protein 1 (TNIP1), ATM Interactor (ATMIN), ST6 N-acetylgalactosaminide alpha-2,6-sialyltransferase 2 (ST6GALNAC2), Neudesin neurotrophic factor (NENF), CCAAT enhancer binding protein zeta (CEBPZ) and leucine rich repeat containing 4 (LARC41) were target of drug Ribavirin (Figure S2). The gene PRKACA can be considered a drug target. PRKACA encodes the catalytic subunit alpha of protein kinase A (PKA), which is an enzyme that plays a role in various cellular signaling pathways, including those involved in metabolism, cell growth, and differentiation. It is reported that there are several drugs that target PKA, either directly or indirectly used in the treatment of certain diseases, such as cancer and genetic mutations in PRKACA have been found in some cases of cortisol-producing adrenal tumors, and drugs that specifically target these mutations are currently being developed and tested in clinical trials^73^. Research has shown that HYOU1 is upregulated in response to various stressors and that it plays a role in protecting cells from stress-induced damage. As a result, HYOU1 has been implicated in the pathogenesis of several diseases, including cancer, cardiovascular disease, and neurodegenerative disorders^74^. Some studies have investigated the potential use of DNAJC15 inhibitors as a therapeutic strategy. For example, one study finds that small molecule inhibitors of DNAJC15 are able to inhibit the growth of cancer cells in vitro and in vivo, suggesting that targeting DNAJC15 can be a promising approach for cancer therapy^75^. There is growing interest in targeting ILK as a potential therapeutic strategy, and several ILK inhibitors have been developed and are currently being evaluated in preclinical and clinical studies^76^. There is some evidence to suggest that TNIP1 may be a potential drug target for inflammation in certain disease models, such as rheumatoid arthritis and lupus. Additionally, small molecule inhibitors of the interaction between TNIP1 and TNFAIP3 have been developed and tested in preclinical models of inflammation^77^. It is reported that CEBPZ promotes the migration and invasion of breast cancer cells and CEBPZ knockdown in liver cancer cells leads to decreased cell proliferation and increases apoptosis^78^.

### Investigation of the potential of the drug to reverse the disease gene expression changes

Disease gene expression reversion has been extensively used to discover new potential treatment for existing drugs^79^. There are a quite large number of up or down regulated gene expressions. We can examine expression reversion one gene at a time. However, such approach has some limitations. First, it overlooks correlation between two gene expressions. Second, it may be infeasible to require that every up or down expressed gene reverses its expressions. Therefore, we constructed networks to represent all disease gene expressions as we showed in the previous section. Then, graph neural networks (GNNs) that use both the graph structure and node features to produce a representation were used for regression.

Specifically, GNNs utilized iterative message passing between neighboring nodes in a network followed by pooling to update node representations and produce node and graph representations. A GNN was composed of multiple layers, with each layer computing representations for all nodes in the network by aggregating information from neighboring nodes and pooling all node representations to generate a network-level representation. The node and network-level representations then were passed to the next layer. After several iterations, the GNN outputed node and network-level representations, which were subsequently used in regression to assess the potential for reversing disease gene expressions.

Initially, we employed GNNs to represent and summarize both directed and undirected networks of gene expressions in response to drug action, as well as directed expression networks that showcase up or down-regulation in disease samples. We then utilized the GNN to summarize the network level of gene expression changes associated with disease, which were subsequently regressed on both drug-directed and undirected perturbation networks. The regression coefficient served as a measure of the potential of the drug to reverse the gene expression changes associated with the disease.

Table 3 presents the results of a regression analysis that examined the potential for four drugs - Chloroquine, Ribavirin, Ritonavir, and Ruxolitinib - to reverse disease gene expressions. Both directed and undirected networks were used to model drug perturbation expression and up/down expression networks in the disease. The results revealed several noteworthy features.

**Table 3.**
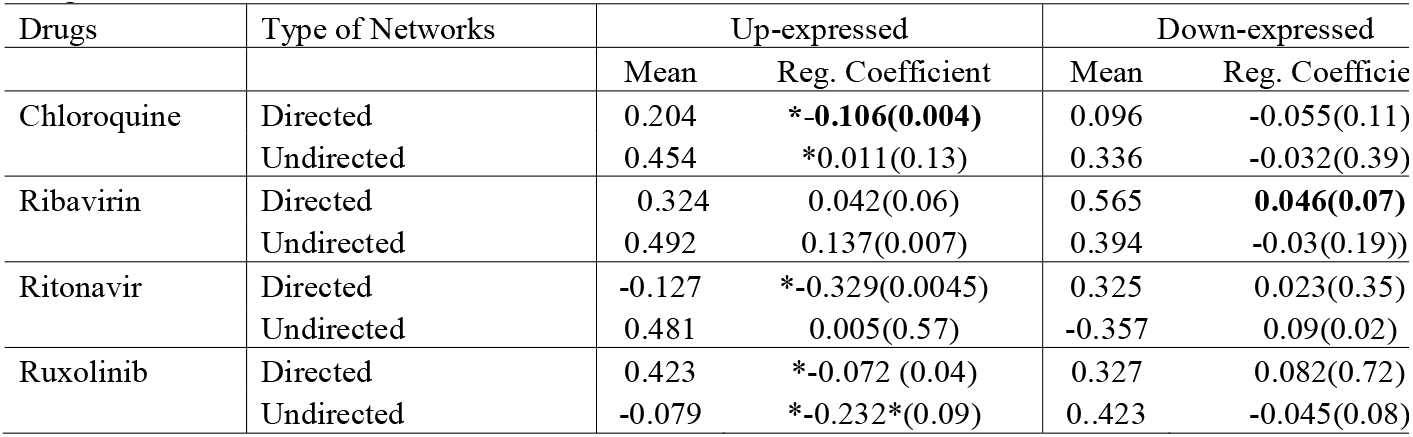
Possibility of reversing up-expressed and down-expressed networks to normal expressions using four drugs.

| Drugs | Type of Networks | Up-expressed |  | Down-expressed |  |
| --- | --- | --- | --- | --- | --- |
|  |  | Mean | Reg. Coefficient | Mean | Reg. Coefficient |
| Chloroquine | Directed | 0.204 | **\*-0.106(0.004)** | 0.096 | -0.055(0.11) |
|  | Undirected | 0.454 | *0.011(0.13) | 0.336 | -0.032(0.39) |
| Ribavirin | Directed | 0.324 | 0.042(0.06) | 0.565 | **0.046(0.07)** |
|  | Undirected | 0.492 | 0.137(0.007) | 0.394 | -0.03(0.19)) |
| Ritonavir | Directed | -0.127 | *-0.329(0.0045) | 0.325 | 0.023(0.35) |
|  | Undirected | 0.481 | 0.005(0.57) | -0.357 | 0.09(0.02) |
| Ruxolitinib | Directed | 0.423 | *-0.072 (0.04) | 0.327 | 0.082(0.72) |
|  | Undirected | -0.079 | *-0.232*(0.09) | 0.423 | -0.045(0.08) |

Firstly, the directed network approach had greater power in detecting the potential effect of reversing disease gene expressions with drugs. The analysis showed a negative coefficient of disease up-expression network regressed on the Chloroquine and Ritonavir directed perturbation network. However, we observed a positive coefficient for the disease up-expression network regressed on the Chloroquine and Ritonavir undirected perturbation networks.

Secondly, the signals indicating the reversal of up-disease expressions were stronger than those indicating the reversal of down-disease expressions. We found significant effects of reversing up-disease expressions using Ritonavir, Chloroquine, and Ruxolitinib, but did not observe significant effects of reversing down-disease expressions using any of the four drugs.

Table 3 presents the estimated effect of reversing the disease expression network. However, it should be noted that this does not guarantee the reversal of disease expressions for all genes. In Table 4, we presented the potential roles of four drugs in reversing the disease expressions of 11 individual genes. Of these genes, seven were up-expressed (DUSP, TICAM1, TP5D1, CCL27, TNFAIP3, TNNT3, CLEC7A), and four were down-expressed (EIF3A, GALNT7, RFX7, IMPACT). While Table 3 showed that three drugs (Chloroquine, Ritonavir, and Ruxolinih) reversed the up-disease expression network, they did not reverse all seven up-expression genes. In contrast, all four drugs showed some potential for reversing either up-expressed or down-expressed genes, as indicated in Table 4. Notably, none of the four drugs showed significant roles in reversing the down-disease expression network, as seen in Table 3. Upon reviewing Table 4, we found that with the exception of CCL27, the disease expressions of all 10 genes were effectively reversed by at least one drug. This suggested that a combination of four drugs might have the potential to reverse the majority of disease expressions. This indicated promising therapeutic potential for reversing the disease-associated gene expressions using drug combinations.

**Table 4.**
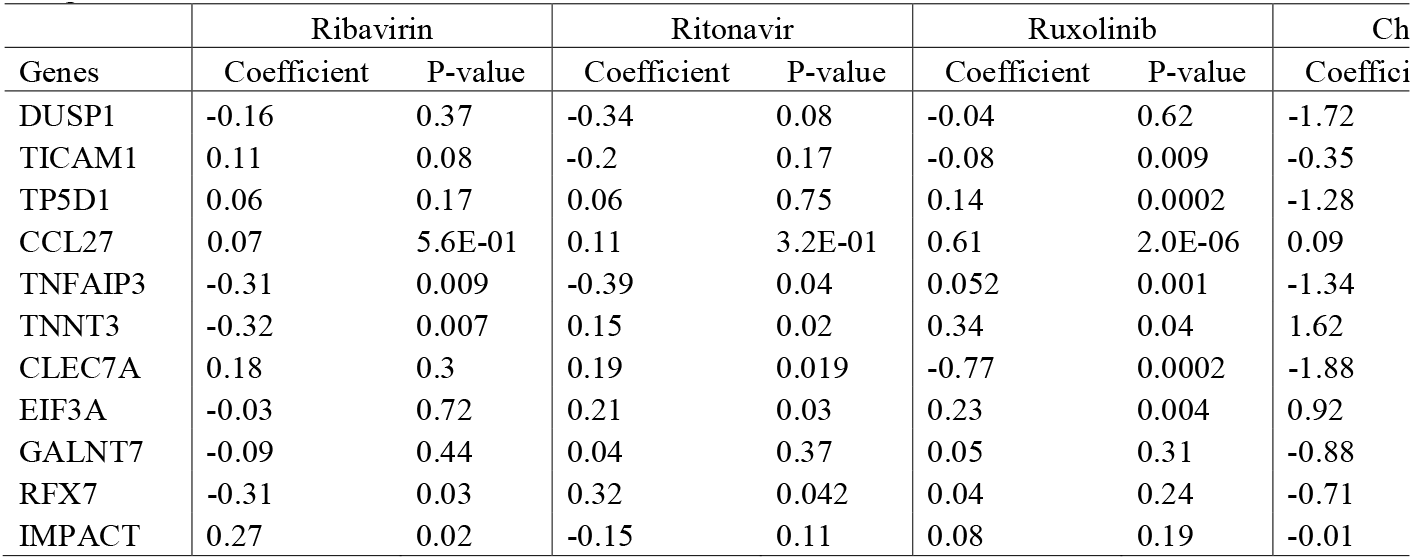
Possibility of reversing over-expressed and under-expressed genes to normal expressions using four drugs.

| Genes | Ribavirin |  | Ritonavir |  | Ruxolinib |  | Ch |
| --- | --- | --- | --- | --- | --- | --- | --- |
|  | Coefficient | P-value | Coefficient | P-value | Coefficient | P-value | Coefficient |
| DUSP1 | -0.16 | 0.37 | -0.34 | 0.08 | -0.04 | 0.62 | -1.72 |
| TICAM1 | 0.11 | 0.08 | -0.2 | 0.17 | -0.08 | 0.009 | -0.35 |
| TP5D1 | 0.06 | 0.17 | 0.06 | 0.75 | 0.14 | 0.0002 | -1.28 |
| CCL27 | 0.07 | 5.6E-01 | 0.11 | 3.2E-01 | 0.61 | 2.0E-06 | 0.09 |
| TNFAIP3 | -0.31 | 0.009 | -0.39 | 0.04 | 0.052 | 0.001 | -1.34 |
| TNNT3 | -0.32 | 0.007 | 0.15 | 0.02 | 0.34 | 0.04 | 1.62 |
| CLEC7A | 0.18 | 0.3 | 0.19 | 0.019 | -0.77 | 0.0002 | -1.88 |
| EIF3A | -0.03 | 0.72 | 0.21 | 0.03 | 0.23 | 0.004 | 0.92 |
| GALNT7 | -0.09 | 0.44 | 0.04 | 0.37 | 0.05 | 0.31 | -0.88 |
| RFX7 | -0.31 | 0.03 | 0.32 | 0.042 | 0.04 | 0.24 | -0.71 |
| IMPACT | 0.27 | 0.02 | -0.15 | 0.11 | 0.08 | 0.19 | -0.01 |

### Target paths in disease up- and down-expression networks

In order to elucidate how drugs can reverse disease-related changes in gene expression, we have provided additional evidence in Figures 6 and 7. Specifically, we showed that administering Ritonavir led to the formation of complex directed networks that were associated with both up- and down-regulated gene expression patterns. Our analysis of these networks, presented in Table 4, revealed that Ritonavir treatment specifically targeted three hub genes, namely DUSP1, TICAM1, and TNFAIP3, in the up-regulated network. Notably, reductions in the expression of these hub genes resulted in the activation of multiple downstream pathways, leading to the formation of highly interconnected and complex networks with 37 genes.

**Figure 6.**
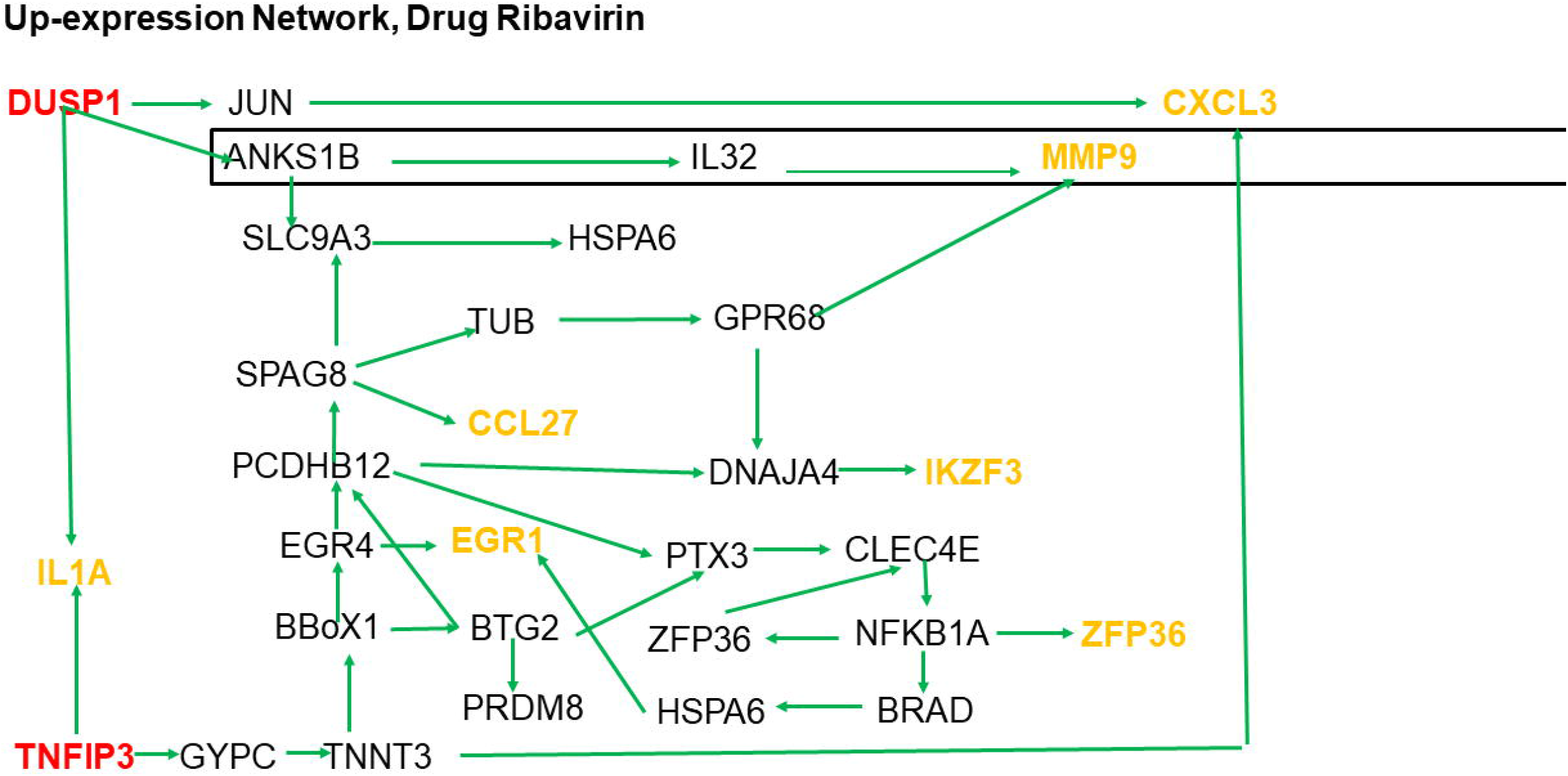
The administration of Ritonavir to reverse disease up-expressions gave rise to intricate directed networks.

**Figure 7.**
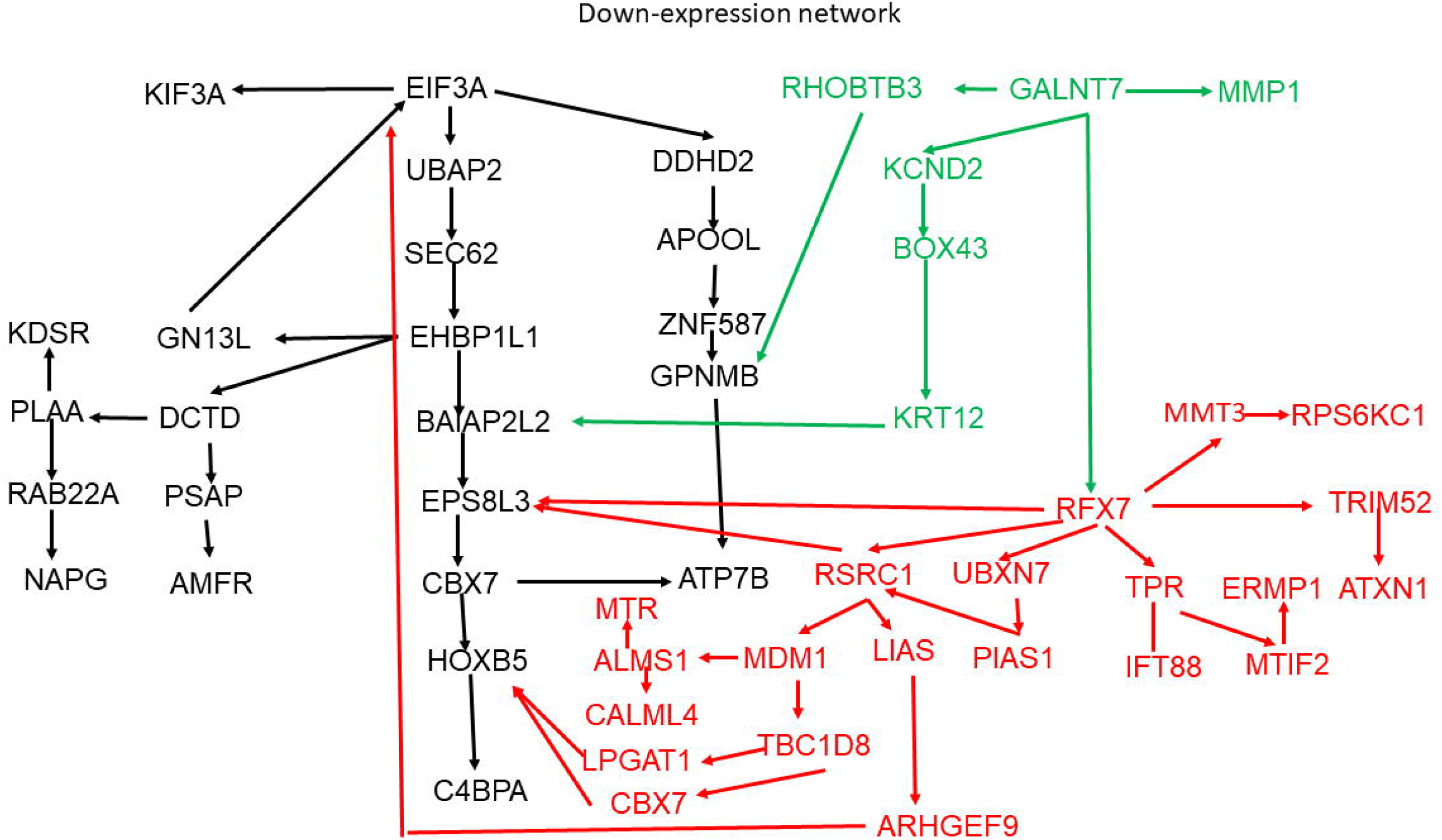
The administration of Ritonavir to reverse disease down-expressions gave rise to intricate directed networks

After reviewing the pathways, we discovered that they led to the following genes: CXCL3, MMP9, IL1A, IKZF3, ZFP36, EGR1, and CCL27. Further research in the literature revealed that each of these genes had been linked to COVID-19.

Studies showed that the CXCL3 gene was involved in the immune response to viral infections, including COVID-19. CXCL3 encoded for a chemokine protein that was produced by various cells of the immune system and was involved in the recruitment of immune cells to the site of infection.

In COVID-19 patients, CXCL3 has been found to be significantly upregulated in the lungs, indicating its potential role in the disease pathogenesis. Additionally, studies have suggested that targeting CXCL3 could be a potential therapeutic strategy for COVID-19, as it could help regulate the immune response and reduce inflammation in the lungs^80^.

There was limited research on the association of the MMP9 gene with COVID-19^81^. The study found that individuals with the mutation had higher levels of MMP9, which was associated with inflammation and lung injury, leading to worse outcomes in COVID-19 patients.

Several studies investigated the association of IL1A gene expression with COVID-19^82^. IL1A might play a role in the pathogenesis of severe COVID-19 and might serve as a potential biomarker for disease severity and prognosis.

ZFP36 was a gene that encoded a protein involved in the regulation of inflammation and immune responses. There had been several studies investigating the association of ZFP36 gene expression with COVID-19^83^. Their findings suggested that ZFP36 might play a role in the regulation of inflammation and immune responses in COVID-19, and might serve as a potential therapeutic target.

EGR1 was a gene that encoded a transcription factor involved in the regulation of cellular growth and differentiation. There had been several studies investigating the association of EGR1 gene expression with COVID-19^84^. EGR1 might play a role in the regulation of immune responses and inflammation in COVID-19, and might serve as a potential therapeutic target.

CCL27 was a chemokine that played a role in the recruitment of immune cells to sites of inflammation. Several studies identified CCL27 as one of the chemokines that was significantly upregulated in the lungs of COVID-19 patients compared to healthy controls^85^.

IKZF3 was a gene that encoded a transcription factor involved in the regulation of immune cell differentiation and function. There was evidence to suggest that other members of the IKZF3 might be involved in the pathogenesis of COVID-19^86^.

Table 4 summarized our analysis of the networks, indicating that Ritonavir treatment specifically targeted three hub genes, EIF3A, GALNT7, and RFX7, in the down-regulated network. Interestingly, upregulation of these hub genes led to the activation of multiple downstream pathways, resulting in the formation of highly interconnected and complex networks involving 50 genes.

In Figure 7, we observed that Ritonavir directly targets MMP1, which is a down-regulated gene in the disease. Additionally, MMP1 expression was found to be associated with the expression of RFX7 (reverse regression coefficient = 0.25 and P-value =0.0004). Furthermore, EGR1 was identified as a targeted gene of Ritonavir and was significantly associated with the expression of RFX7 (reverse regression coefficient = 0.56 and P-value = 0.00005), as shown in Figure 7. Three genes, GALNT7, MMP1 and EGR1 transcriptionally regulated RFX7, which in turn regulated 25 genes in the down-regulated expression network in COVID-19.

RFX7 is transcription factor. It is reported that RFX7 may also have a role in neurological and metabolic disorders. It limits metabolism of NK cells and promotes their maintenance and immunity^87^. A Multi-omics analysis identifies RFX7 targets involved in tumor suppression and neuronal processes^88^.

MMP1 is a drug target gene^89^. There are several drugs that target MMP1 for the treatment of various diseases, including small molecule inhibitors and monoclonal antibodies. EGR1 is also a drug target gene. EGR1 is a transcription factor that plays a role in the regulation of cell growth, differentiation, and apoptosis, and its dysregulation has been implicated in various diseases, including cancer, cardiovascular disease, and inflammatory disorders. There are several drugs in development that target this gene for the treatment of various diseases^90^.

Further analysis revealed that paths originating from the three hub genes, EIF3A, GALNT7, and RFX7, in the down-regulated network were reversed in the disease state. Remarkably, these paths eventually converged on gene C4BPA.

There have been some studies investigating the association between C4BPA and COVID-19.

Overall, several studies suggest that C4BPA may play a role in the immune response to SARS-CoV-2 and that genetic variations in the C4BPA gene may influence susceptibility to severe COVID-19^91^.

## Discussion

In this study, we have proposed a novel approach for identifying potential drug repurposing candidates that utilizes a combination of graph learning and causation analysis. A key challenge in conducting causal inference in drug repurposing studies is developing a robust and large-scale causal network (DAG) with thousands of nodes. However, the classical methods for inferring DAGs are often limited by a discrete optimization problem that requires an extensive search, resulting in an intractable search space that seriously limits the size of the reconstructed DAG.

To overcome the limitations of combinatorial optimization, we have formulated DAG learning as a continuous optimization problem with acyclic constraints. Through large-scale simulations, we have demonstrated that our proposed method for DAG learning is robust and has high power in reconstructing DAGs with up to 1,000 nodes.

To illustrate that our approach has significant potential for identifying drug repurposing candidates, the proposed methods were applied to L1000 perturbational profiles of 978 genes from Broad Institute LINCS center under 4 drugs: ritonavir, chloroquine, ruxolinib and ribavirin to reconstruct drug induced causal networks, and to up- and down-gene expressions in post-mortem lung samples from COVID-19-positive patients to reconstruct disease causal expression networks. To our knowledge, this has been the first time to construct causal networks with up to 1,000 nodes.

The causal network approach is a powerful tool for identifying potential drug repurposing candidates. Here are some advantages of using this approach:

1. Uncovering causal relationships: Causal network analysis can reveal causal relationships between different genes, proteins, and pathways, providing insights into the mechanisms underlying disease. We have showed that administering Ritonavir leads to the formation of complex directed networks with 85 genes that were associated with up- or down-regulated gene expression patterns.
2. DAG analysis revealed that Ritonavir reversed disease up- and down-gene expressions to normal level, but the undirected network analysis completely failed.
3. By identifying the causal factors that contribute to the disease, the approach can help identify new drug targets and repurposing candidates. Causal network analysis elucidated that how drugs reversed disease-related changes in gene expression. It revealed that Ritonavir treatment specifically targeted three hub genes, namely DUSP1, TICAM1, and TNFAIP3, in the up-regulated network. After reviewing the pathways, we discovered that they lead to the following genes: CXCL3, MMP9, IL1A, IKZF3, ZFP36, EGR1, and CCL27. Further research in the literature revealed that each of these genes has been linked to COVID-19. Causal network analysis also found that Ritonavir treatment specifically targeted three hub genes, EIF3A, GALNT7, and RFX7, in the down-regulated network, which lead to three drug target genes: MMP1, EGR1 and C4BPA.
4. Prediction of drug efficacy: The causal network approach can predict the efficacy of a drug by identifying the causal relationships between drug targets and disease pathways. This can help prioritize repurposing candidates for further investigation.
5. Reducing cost and time: Repurposing drugs can save time and cost compared to developing new drugs. The causal network approach can help identify repurposing candidates with a higher likelihood of success, reducing the time and cost of drug development.

To investigate the potential of the causal network approach for identification of drug repurposing candidates, we examined four drugs: Ritonavir, Chloroquine, Ribavirin, and Ruxolitinib. Our results showed that reversing up-disease expressions using Ritonavir, Chloroquine, and Ruxolitinib had significant effects, while we did not observe significant effects of reversing down-disease expressions using any of the four drugs. Additionally, we found that Ritonavir was slightly more likely to be a better candidate for treating COVID-19 than the other three drugs. However, due to the small sample size of publicly available Nirmatrelvir induced gene expression data, we were unable to estimate the effects of reversing disease gene expressions to a normal level using Nirmatrelvir and Paxlovid.

In this study, GNNs are used to investigate the potential for reversing disease gene expressions. They have several advantagesfor investigating such potential:

### Advantages

1. Capturing complex relationships: GNNs can capture complex and non-linear relationships between genes and their expressions, making them useful for modeling the complex interactions that occur in gene expressions and drug responses.
2. Integration of multiple data types: GNNs can integrate multiple types of data to provide a more comprehensive view of disease mechanisms.
3. Predictive power: GNNs have demonstrated excellent predictive power for various tasks, including drug discovery and predicting gene-disease associations.
4. Scalability: GNNs can be scaled to handle large datasets, making them suitable for analyzing large-scale biological data.

However, there are also some limitations to using GNNs for investigating the potential for reversing disease gene expressions:

### Limitations

1. Interpretability: The complex structure of GNNs makes it challenging to interpret the model’s results and understand the underlying mechanisms.
2. Although GNNs reveal that disease up- or down-expressions are reversed at the network level, it does not necessarily mean that the disease expressions of all genes are restored to normal levels. Our observation shows that Ritonavir reversed the disease up-expression network, but it only restored three disease gene expressions out of the seven hub genes in the disease up-expression network.
3. Generalization: GNNs may have difficulty generalizing to new datasets or diseases that have different biological mechanisms or genetic backgrounds.
4. Biased representation: GNNs can be biased towards the data used for training, which may lead to overfitting or inaccurate predictions when applied to new datasets.

Overall, while GNNs have several advantages for investigating the potential for reversing disease gene expressions, it is essential to carefully consider their limitations and ensure that their results are validated by experimental studies.

Our study has revealed that none of the four drugs tested were able to reverse all 11 hub genes in the up- and down-expression networks associated with the disease. However, with the exception of one gene, all 10 other genes showed significant improvement in disease expression after treatment with at least one of the drugs. This suggests that a combination of the four drugs may be able to effectively reverse the majority of disease expressions. These findings indicate that the proposed approach allows to find a promising therapeutic strategy for addressing the disease-associated gene expressions using drug combinations.

Repurposing drugs for treating diseases presents a significant challenge due to the complex networks formed by a large number of pathways between drug targets and disease genes. Although we have developed an approach to reconstruct causal networks up to 1,000 nodes, we still lack efficient methods for integrating multimodal data from genomic, transcriptomic, epigenetic, proteomic, and metabolic sources across multiple cell types. Furthermore, reconstructing large-scale cascade DAGs with thousands of nodes, starting from drug targets through multiple directed paths (transcriptomic, epigenetic, proteomic, metabolic) and ending with diseases, remains a challenge. One of the most significant limitations of the drug repurposing approach is the lack of a bridge between the drug signature and the reverse disease signature. While the use of rapidly developing artificial intelligence tools and molecular biology technologies shows promise for developing a general and efficient framework for identifying drug repurposing candidates, we still have a long way to go.

## Methods

### Structural equation model

Let *Y* = [*Y*_1_,…,*Y*_*M*_]^*T*^ be a vector of the *M* endogenous variables, e.g., gene expressions and *X* = [*X*_1_,…, *X*_*K*_]^T^ be a vector of *K* exogenous variables, e.g., drug dosage. We denote the errors by *e*. We assume that *E*[*e*] = 0 and that *e* is uncorrelated with the exogenous variables in *X*. We also assume that *e*_i_ is homoscedastic and nonautocorrelated^20^. Assume that the sample size is *n*, the number of endogenous variables is *M*, and the number of exogenous variables is *K*. Then, structural equation model (SEM) are given by

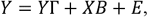

where

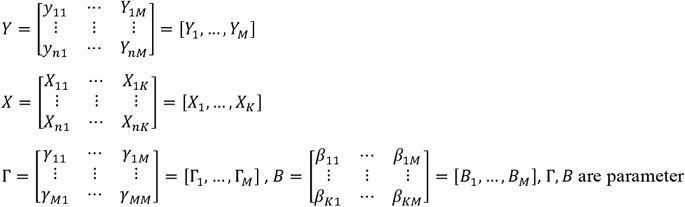

matrices, and

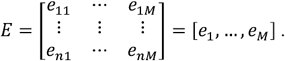

### Parameter estimation and optimization in the SEM

After introducing the matrix exponential as a constraint to ensure the network’s acyclicity, the optimization problem for learning DAGs using SEMs are reduced to

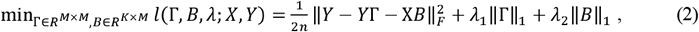

Subject to h(Γ) = Tr(*e*^Γ∘Γ^) − *M* = 0, where ∥·∥_*F*_ denotes frobenius matrix norm, ∥·∥_1_ denotes *L*_1_ norm and ∘ denotes an element multiplication.

Now the combinatorial optimization problem is transformed to equality constrained continuous, but nonsmooth optimization problem. Solutions consists of two major steps: (1) using Lagrange multiplier method to transform the equality constrained optimization problem into a sequence of non-constrained optimization problem and (2) solving non-smooth optimization problem.

### Transform the Equality Constrained Optimization Problem into Unconstrained Optimization Problem

The classical augmented Lagrange multiple method is employed to convert the constrained optimization (2) into an unconstrained optimization problem:

The primal problem:

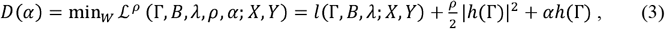

where

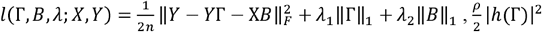 is an augmentation term and *αh*(Γ) is a Lagrange term.

The dual problem:

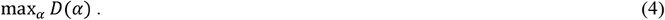

The optimization problem (3) can be separated into smooth optimization and nonsmooth optimization problem:

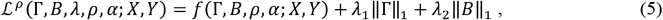

where

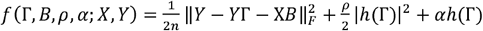

is the smooth part of the objective function. For simplicity of notation, we define 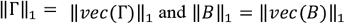 and Let 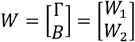 and *ω* = *vec*(*W*) Then, equation (5) can be rewritten as

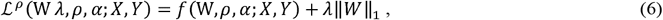

where

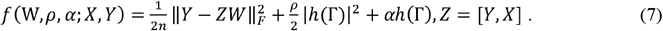

Newton’s classic method is an efficient approach for solving unconstrained smooth optimization problems, but it is not suitable for nonsmooth optimization problems. To address this, we will use proximal methods, which can be seen as an extension of Newton’s method that enables us to solve nonsmooth optimization problems^92^. In general, the optimization problem (5) can be solved by the proximal method^92,93^. In other words, to solve the optimization problem (5), at each iteration we often expand the function *f*(Γ,*B,ρ,α,X,Y*) in a neighborhood of the current iterate (Γ_*i*_,*B*_*i*_) by a Taylor expansion:

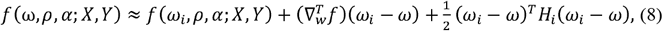

where

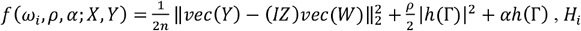, *H*_*i*_ is the Hessian matrix and *ω* is viewed as *vec*(*W*).

The Hessian matrix *H*_*i*_ can be approximated by BFGS correction^94^. We can show that BFGS correction *E*_*i*_ is given by,

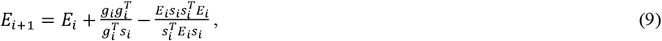

where

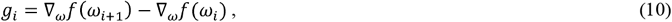

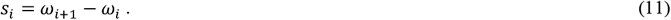

Thus, equation (8) can be reduced to

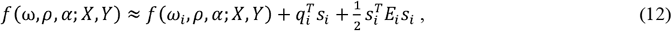

where *q*_i_ = ∇_*ω*_*f*(*ω*_*i*_).

Combining equations (5) and (12), optimization problem (5) at iteration *i* can be reduced to

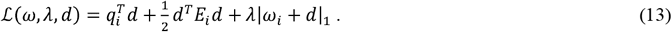

Coordinate descent algorithms, which solve optimization problems by successively perform optimization along coordinate directions and often have a closed form solution, can be used to solve nonsmooth optimization problem (13). Let *e*_*j*_ be an unit vector with the *j*^*th*^ component being 1 and all other components being zero. For simplicity, the changes of the *j*^*th*^ component of *W*_*i*_ + *d* is denoted by *W*_*j*_ + *d*_*j*_ + *ze*_*i*_ where *z* is a search variable. For the *j*^*th*^ component, equation (13) is reduced to

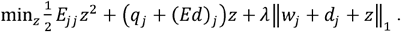

The solution is

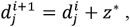

where

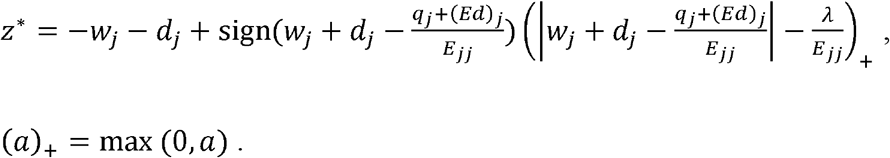

### Graph neural networks provide a general and flexible framework for describing and analyzing network data

Graph neural networks (GNNs) provide a condensed representation of an input graph. The key insight behind GNNs is that the representation of a node in a graph should be influenced by the representations of its neighbors. This is achieved through a message-passing scheme, where each node aggregates information from its neighbors, updates its own representation, and then sends a message to its neighbors.

Consider a graph *G* = (*V,E*), where *V* denotes *n* nodes and *E* = (*e*_*ij*_) denotes a set of edges. Let 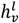 denote an embedding of node in the layer. Learning GNN is implemented by the following iterative algorithm:

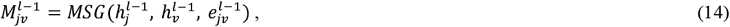

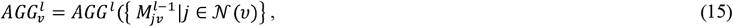

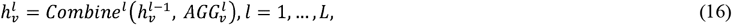

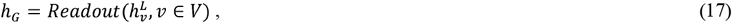

where 𝒩(*v*) denotes a neighborhood of the node *v*, MSG, AGG, Combine and Readout are implemented by feedforward neural networks.

### Directed acyclic graph neural networks

Directed Acyclic Graph Neural Networks (DAGNNs) are a type of neural network that can model and analyze complex data structures with a directed acyclic graph. Algorithm for updating node representations is based on those of all their predecessors sequentially, such that nodes without successors digest the information of the entire graph. The DAGNN learning algorithm is given by^20^

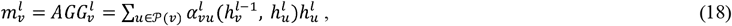

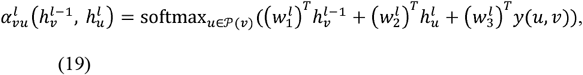

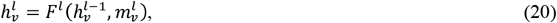

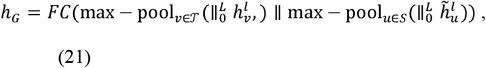

where *P*(*v*) denotes the set of direct predecessors of *v, T* denotes the set of nodes without (direct) successors, 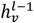 is defined as a query, 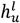 is defined as a key, or a value, the weighting coefficients 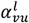 are defined by the standard attention mechanism and calculated by equation (19), 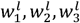 are model parameters, *y*(*u,v*) are representation of edge (*u,v*), *F*^*l*^ is the combine operator that combines the message 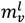 with the previous representation of *v*, 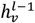, and *FC* represents a fully-connected layer.

Learning algorithm can be a bidirectional process. The directions of the edges in G can be optionally inverted to create a reverse *DAG* 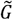. The representation of node *v* in 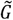 at the *l*-th layer is denoted by 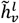. After *L* layers of (bidirectional) processing, the graph level representation is generated by the computed node representations. The representations across layers are concatenated, a max-pooling across nodes are performed, and a fully-connected layer is applied to produce the output *h*_*G*_^20^.

### Graph regression

Regression can be extended to graph regression where both sides of regression are graph representations. The GNNs and DAGNN can be used to summarize information of undirected graph and DAG. Suppose that we regress one graph on another graph. Let *y* be representation of the graph at the graph level and *x* be representation of another graph at the graph level. The representation can be a single value or vectors of values.

Let’s consider the regression of one graph on another graph. Let *y* be the representation of the graph at the graph level, and *x* be the representation of another graph at the graph level. These representations can be a single value or vectors of values, depending on the chosen approach. Graph regression allows us to model the relationship between graph-level representations, enabling tasks such as prediction, inference, and analysis across graphs.

Define a regression:

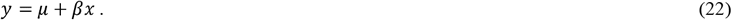

Then, the regression (22) that uses representations of graphs as dependent variable and independent variables is referred as graph regression.

## Data availability

All datasets used in this work are publicly available from the following sources: The gene expression data for SARS-CoV-2 were obtained from GSE147507 (https://www.ncbi.nlm.nih.gov/geo/query/acc.cgi?acc=GSE147507). The CMap and L1000 data were downloaded from GSE 92742 and GSE 70138 (perturbational profiles from Broad Institute LINCS center, phase I and II) (https://www.ncbi.nlm.nih.gov/geo/query/acc.cgi?acc=GSE92742 https://www.ncbi.nlm.nih.gov/geo/query/acc.cgi?acc=GSE70138).

## Code availability

The code for reconstruction of DAG is submitted to github https://github.com/SplinterTao/DAG. We relied on open source libraries for reconstruction of undirected graph and implementations of GNN and DAGNN.

## Acknowledgments

Grant supported information

Drs. Jinying Zhao and Tao Xu are supported by NIH grants R01Dk107532 and 7RF1AG052476.

## Author Contribution Statement

Perform data analysis: Tao Xu

Provide financial support for Dr. Xu and design project: Jinying Zhao

Design project and write a paper: Momiao Xiong

## Figure S Legend

**Figure S1**. Network of gene expressions induced by Ruxolitinib with causal implications.

**Figure S2**. Network of gene expressions induced by Ribavirin with causal implications.

