## Supplemental Figure S1 and Figure S2 for "Graphical Learning and Causal Inference for Drug Repurposing": Supplemental Files.docx

**Figure S1:** Network of gene expressions induced by Ruxolitinib with causal implications.

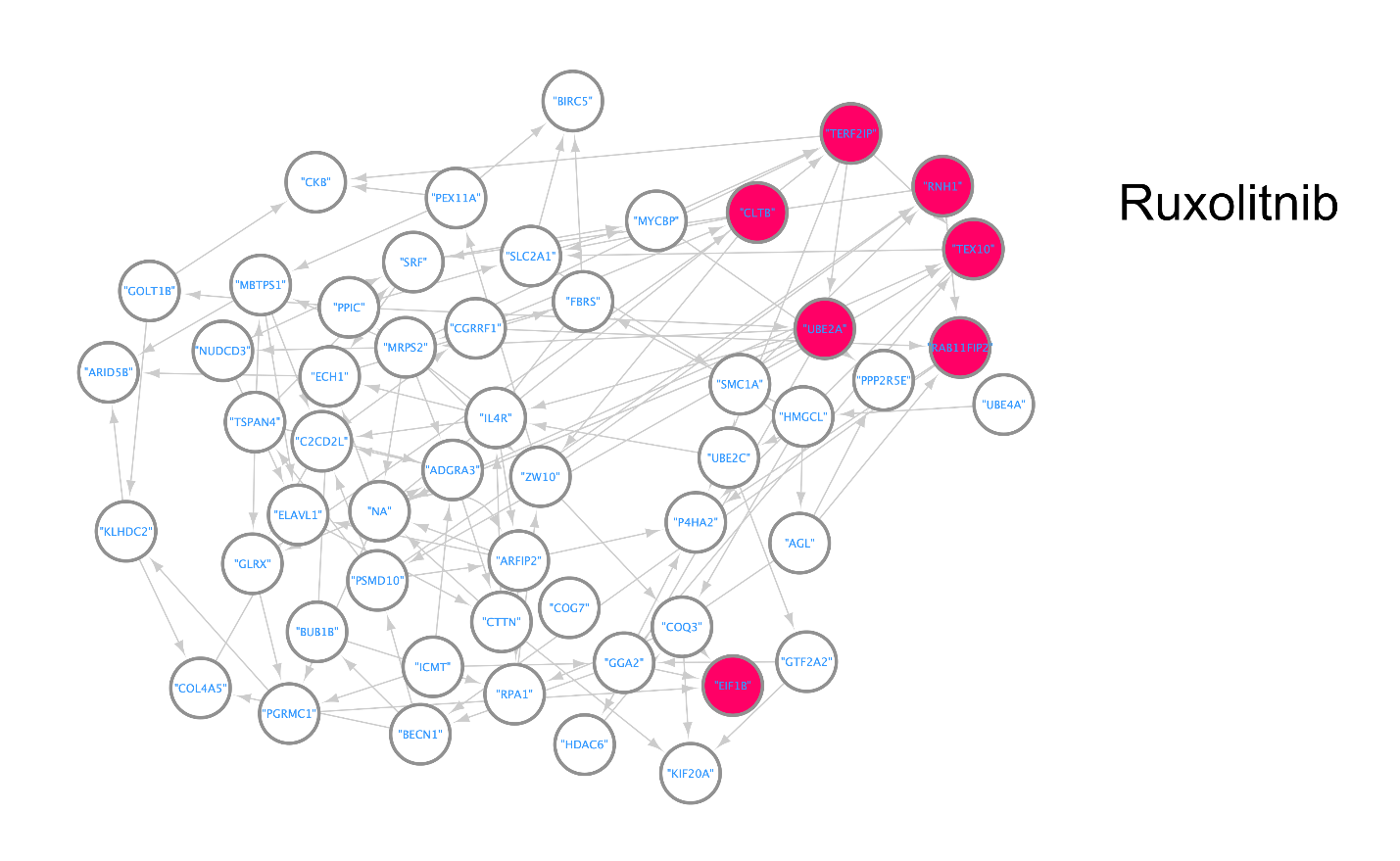

**Figure S2.** Network of gene expressions induced by Ribavirin with causal implications.

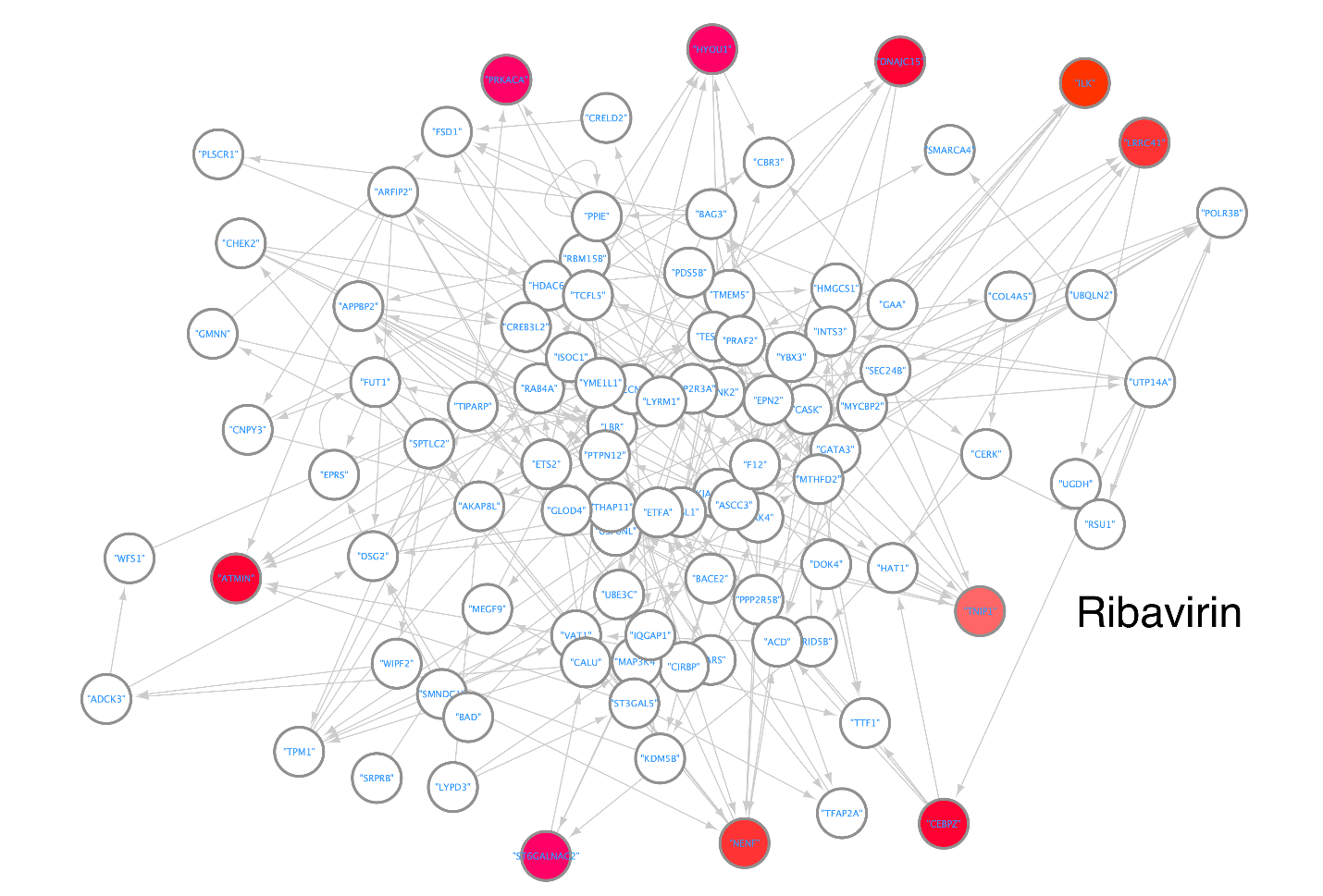
